# Linkage between HLA-B8 and HLA-DQ2.5 Contributes to Ancestry-Dependent Genetic Risk for Celiac Disease

**DOI:** 10.1101/2024.12.20.24319436

**Authors:** Xin Long, Hemanth Karnati, Wenjing Ying, Mary-Joe Touma, Ioana Smith, Suzanne Lewis, Chao Xing, Ezra Burstein, Peter HR Green, Michele J. Alkalay, Alexandre Bolze, Xiao-Fei Kong

**Author notes:** Correspondence: Xiao-Fei Kong, MD, Ph.D, Division of Digestive and Liver Diseases, Dept of Internal Medicine McDermott Center for Human Growth and Development, UT Southwestern Medical Center, 5323 Harry Hines Blvd, Suite J5.150 Dallas, TX, 75390-9151. **Author Contributions:** All authors provided important insights in the analysis and interpretation of the data and were involved in editing the manuscript. X.F.K. wrote the manuscript. **Disclosures:** The authors have no relevant financial or non-financial interests to disclose. **Statement:** The data and code used in this study are available as a shared workspace to registered researchers of the *All of Us* Researcher Workbench. For information about access, please visit https://www.researchallofus.org/. Computational codes are available from: https://github.com/Kong-Lab-UTSW/.

## Abstract

**Background:** Most genetic studies on celiac disease (CeD) have focused on individuals of European descent. Limited data are available for the Hispanic and black populations.

**Methods:** We analyzed whole-genome sequencing data, electronic health records (EHR), and laboratory results from the *All of Us* Research Program. We identified 3,481 individuals with CeD through EHR, self-reporting, or both. Of these, 2,899 carried one of the four well-established risk haplotypes, including 262 of admixed American (89% Hispanic) and 108 of African (70% black) ancestry. Five sex-, age-, and ancestry-matched controls per case were selected for the assessment of genetic and clinical risk factors.

**Results:** An enrichment in the *DQB1*02:01* allele was observed in CeD patients across all ancestries, with the strongest association in Europeans (32.3% vs. 11.6%), followed by Americans (18.5% vs. 8.1%) and Africans (15.7% vs. 8.1%). Among individuals carrying the DQ2.5 (*DQA1*05:01-DQB1*02:01* haplotype), HLA-B8 was present in 72.3% of Europeans, 42.3% of Admixed Americans, and lower in Africans. This linkage disequilibrium was higher in CeD patients than in controls across all three ancestries. A polygenic risk score distinguished seropositive CeD from controls with 86% accuracy.

Incorporating clinical risk factors, including family history, hypothyroidism, diarrhea, vitamin D deficiency, and anemia, increased predictive accuracy to 92%. The model identified 93% of CeD patients with tTG-IgA levels greater than 10 IU/mL.

**Conclusion:** Linkage between HLA-B8 and DQ2.5 differs significantly among individuals of European, admixed American, and African ancestry, contributing to ancestry-dependent genetic risk for CeD.

**Summary:** Using data from the *All of Us Research Program*, we found that HLA-DQ2.5, a key genetic risk factor for celiac disease, is present in European (11.6%), Admixed American (8.1%), and African (8.1%) populations, but its disease penetrance differs significantly by ancestry.

## Introduction

Celiac disease (CeD) has a worldwide prevalence of about 1%, ranging from 0.7 to 2%, depending on the tools used for screening and the country(1,2). However, most CeD cases remain undiagnosed(3). Clinically, the recognition of CeD early in life would be beneficial to patients because undiagnosed CeD can lead to various complications, including osteoporosis, iron deficiency anemia, poor quality of life, and even cancer(4,5). The only treatment available for CeD is adherence to a gluten-free diet (GFD), which is not always effective, as only 67% of individuals achieve mucosal healing after five years(6). Recent advances have led to the development of additional nomenclatures for describing the various phenotypes related to CeD, such as seronegative CeD, potential CeD, and refractory CeD, further complicating the diagnosis of this condition(7–9). The genetic basis of this clinical variability remains unclear, and the optimal clinical management for these phenotypes remains to be determined(10).

Human genetics plays a key role in the development of CeD. Twin studies have shown that genetic factors account for 70% of the overall risk of CeD(11,12), with CeD occurring in 10% of the first-degree relatives of index cases(13). *HLA-DQA1* and *HLA-DQB1* are two adjacent genes that encode the alpha and beta chains, respectively, to form HLA-DQ heterodimers. The four CeD-compatible HLA risk heterodimers encoded by four haplotypes include DQ2.5 (*DQA1*05:01*-*DQB1*02:01*), DQ2.2 (*DQA1*02:01*-*DQB1*02:02*), DQ8.1 (*DQA1*03:01*-*DQB1*03:02*), and DQ7.5 (*DQA1*05:05-DQB1*03:01*)(14), for which testing is recommended by current guidelines in various clinical scenarios (15,16). Throughout the manuscript, we use standard clinical shorthand (e.g., ‘DQ2.5’) to denote HLA-DQ heterodimers. These are not gene symbols but heterodimers designations referring to specific *DQA1*/*DQB1* allele combinations, as defined by the WHO HLA nomenclature. HLA-DQ2.5 has made a major contribution to our understanding of the pathogenesis of CeD in recent decades, as 95% of European CeD patients carry HLA-DQ2.5 in a *cis* or *trans* configuration (17,18). However, most of these studies were performed in Europe and included participants from the United Kingdom, Italy, and the Netherlands, with very few individuals of other ancestries represented(18,19). This lack of ethnic diversity in genetic studies has limited our understanding of disease risk across different genetic backgrounds. *All of Us,* a large population-based genetic study, has enrolled more than 700,000 individuals from the United States, 46% of whom belong to underrepresented racial and minority ethnic groups (20). This NIH-supported project provides an unprecedented opportunity for advancing disease prevention and treatment and enhancing diversity in medical studies (20,21). We therefore made used of the *All of Us* datasets to investigate ancestry-specific genetic risks for CeD and to evaluate a risk score that integrates genetic variants, including HLA genotypes, along with clinical risk factors for the prediction of CeD risk across diverse populations.

## Methods

### Ethics Statement

The Ethics Committee/Institutional Review Board (IRB) of the *All of Us* Research Program (AoURP) gave its approval for the collection and analysis of data from human participants (AoU IRB Protocol Number: 2021-02-TN-001). This study was approved by the *All of Us* Research Program Science Committee. The results are reported in accordance with the *All of Us* Data and Statistics Dissemination Policy and are displayed only for groups of at least 20 individuals.

### Data sources

The AoURP aims to build one of the largest and most diverse health databases by enrolling one million adults and children from all backgrounds who have consented to participate and share their electronic health records (EHRs). These records include inpatient and outpatient visits, international classification of diseases (ICD) diagnostic codes, physician notes, laboratory results, and more(20,22–24). Interested participants complete a basic survey. Numerous institutions across the United States are collaborating with *All of Us* to facilitate in-person visits, serving as enrollment centers where individuals can provide physical measurements and biological samples.

We analyzed controlled-tier datasets from the Curated Data Repository (CDR, C2024Q3R4 version), which includes survey responses, EHRs, measurements, and genomic data from participants enrolled between May 31, 2017, and Oct 01, 2023. Whole-genome sequencing (WGS) data were available for 414,830 participants (Fig. 1A), including compressed reference-oriented alignment map (CRAM) files and variant data store (VDS) files. Variant analysis was performed with the Hail framework. The control group consisted of participants without CeD. Individuals with autoimmune thyroid disorders, type 1 diabetes (T1D), selective IgA deficiency, inflammatory bowel disease (IBD), HIV infection, common variable immunodeficiency, cancer, Down syndrome, or DiGeorge syndrome were excluded from the control group. Genetic ancestry was inferred from SNP data by principal component analysis. Propensity score matching based on age, sex and genetic ancestry was performed to identify appropriate controls. Tissue transglutaminase 2 (tTG) and deamidated gliadin peptide (DGP) antibody levels (tTG-IgA, DGP-IgA, and DGP-IgG) were analyzed to identify participants with seropositive CeD. Because those measurements can vary across laboratories and reagents, the cutoffs differ accordingly. Although the specific tTG-IgA assay kits used for these participants were not available, individuals with a reported positive result or with tTG-IgA, DGP-IgA, or DGP-IgG levels ≥20 IU/mL were classified as seropositive.

**Figure 1.**
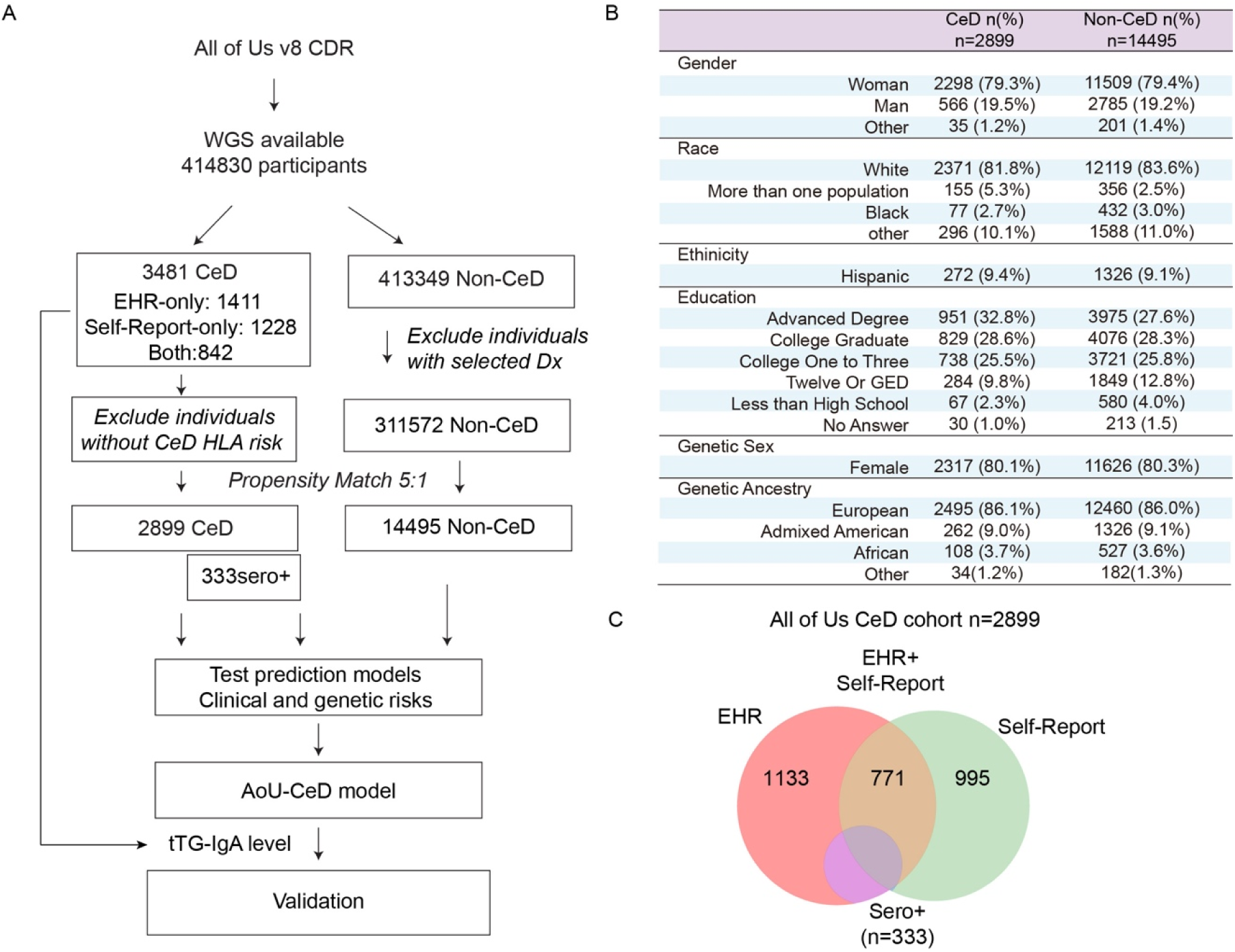
CONSORT flow diagram and cohort demographics for CeD analysis. (A) CONSORT-style flow diagram illustrating the steps of data processing and cohort selection. Propensity score matching was performed based on age, sex, and genetic ancestry. Confounding was reduced by excluding individuals with comorbidities commonly associated with CeD, including T1D, autoimmune thyroid disorders, selective IgA deficiency, IBD, CVID, cancer, and HIV infection, from the non-CeD control group. (B) Demographic characteristics of 5:1 propensity matched Non-CeD controls and CeD patients from *All of Us*. GED, General Educational Development. (C) Venn diagram showing the route of CeD diagnosis for the included participants. “Seropositive CeD” refers to individuals with tTG-IgA or DGP IgA or IgG levels >20 IU/mL documented in their EHRs. (D) Age, sex, and genetic ancestry of the 1:5 propensity score–matched CeD cases and controls.

### Ancestry-specific accuracy of HLA imputation

Participants with potential celiac disease, seronegative celiac disease, gluten intolerance, or non-celiac gluten sensitivity may have been labeled as CeD. To better characterize the cohort, we performed HLA typing using genomic data (20). We tested three approaches on 794 samples to obtain a fast accurate method for HLA typing: HLA genotype imputation with attribute bagging (HIBAG)(25), HLA*LA(26) and the tagSNPs(27) approach (S.Fig1).

We first investigated ancestry-specific HLA imputation accuracy by comparing tagSNP-based inference with HIBAG. One commonly used tagSNP, rs2187668 (27), is widely applied to infer HLA-DQ2.5 and showed excellent concordance in participants of European and African ancestry (98.5% and 97.3%, respectively), but substantially lower concordance in those of admixed American ancestry (62.7%, S.Table2). Consistent with the prior report (27), tagSNPs perform well for DQ7.5 and DQ8.1 but are suboptimal for DQ2.2 in individuals of European ancestry. Concordance between tagSNP and HIBAG was lower in participants of non-European ancestry (S.Table2).

HIBAG is a machine learning-based imputation tool that uses ancestry-specific, pre-trained classifiers to predict HLA genotypes from SNP data (28), whereas HLA-LA is a graph-based method that uses all the HLA reference sequences from the IMGT/HLA database to enable accurate high-resolution typing across diverse ancestries (26). For HIBAG, genetic data from subjects of European (*n*=2668), Asian (*n*=720), Hispanic (*n*=439) and African (*n*=173) ancestries were used in a machine-learning approach to develop accurate HLA prediction. With WGS data from 797 participants, concordance between HIBAG and HLA-LA exceeded 90% at the *HLA*-*A*, *B*, *C*, *DQB1*, and *DRB1* loci (S. Fig. 1). The largest discrepancy was observed for *HLA-DQA1,* largely reflecting reduced performance in distinguishing *DQA1*05:01 and DQA1*05:05*. We then leveraged known linkage disequilibrium (LD) among *HLA-DQA1*, *HLA-DQB1*, and *HLA-DRB1* to evaluate internal consistency. For the DQ2.5, *DQA1*05:01–DQB1*01:01–DRB1*03:01* linkage was strong (>99%) across individuals of European and admixed American ancestry (Fig 2A). In contrast, individuals with African ancestry showed lower linkage: among 119 *DQB1*02:01* alleles, 103(86.6%) were linked to *DQA1*05:01* (Fig 2A). This pattern suggests greater allelic diversity at the *HLA-DQA1* locus in African populations, which may also contribute to reduced *DQA1* concordance. We checked the internal consistency of the HLA-DQ haplotype by applying ancestry-specific linkage disequilibrium to *HLA-DQB1/DRB1*(29). The most common three-locus haplotypes are provided in the Supplementary Table3. Because HIBAG is cost-effective and supports ancestry-specific classifiers, whereas HLA-LA provides superior accuracy at a higher computational cost, we used HIBAG for HLA typing in the full cohort.

**Figure 2.**
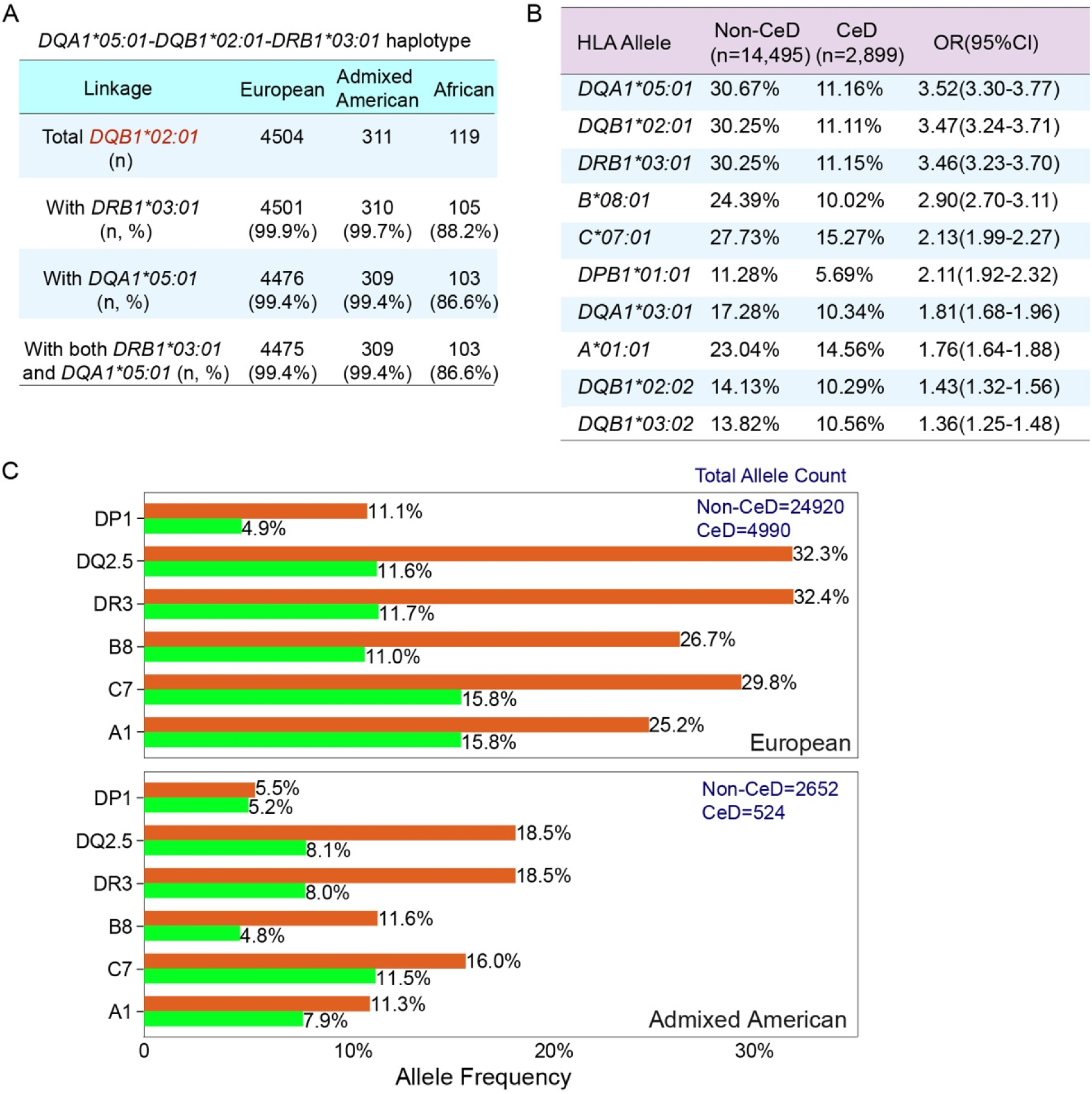
HLA alleles and linkage disequilibrium patterns with DQ2.5 in CeD patients and matched controls across ancestries. (A) Cross-ancestry LD between HLA-DQA1, HLA-DQB1, and HLA-DRB1 in DQ2.5 carriers. (B) Table showing HLA alleles from the HLA-*A,-B,-C,-DPB1,-DQA1,-DQB1*, and - *DRB1* loci overrepresented in individuals with CeD, with a frequency at least 1% higher in such individuals than in matched controls. (C) Allele frequencies for HLA-DQ2.5 and other alleles in linkage disequilibrium with HLA-DQ2.5 (A1, B8, C7, DRB3 and DPB1) across different genetic ancestries and CeD statuses.

## Statistical analysis

We evaluated genetic differences between the CeD and non-CeD participants, regardless of genetic ancestry and HLA-DQ risk genotypes and taking these factors into account. Chi-squared tests were performed for categorical variables. ANOVA was performed to compare differences in numerical variables across more than three groups, followed by Tukey’s honestly significant difference post-hoc test to identify group means and their corresponding 95% confidence intervals. False discovery rate (FDR) correction was applied to adjust for multiple comparisons. Logistic regression analyses were performed to identify independent genetic risks and comorbid conditions, with age, sex, and the principal components of genetic ancestry as covariates. The prediction model was developed with a logistic regression framework combined with machine learning techniques to identify the optimal features. All analyses were conducted within the *All of Us* Researcher Workbench, a cloud-based platform providing data access(20,22). The analysis was performed with Python and the pandas, statsmodels, numpy, scikit-learn, matplotlib, and seaborn packages.

## Results

### Characteristics of participants with celiac disease in All of Us Research Program

The AoURP collaborates with hospitals, commercial laboratory networks, and community organizations to enroll one million U.S. residents across diverse racial and ethnic backgrounds. While the program is designed to be inclusive and now provides WGS data for 414,830 participants, not all participants completed the surveys. Because electronic health record (EHRs) systems vary widely across the nation, not all participants have EHRs or laboratory data. We identified 3,481 of these participants as having CeD (Fig. 1A, S. Table 1) based on the diagnosis in their EHRs or self-report in surveys. As an exploratory cohort, 78% were female and 86% were white, both proportions being significantly higher than in the rest of the cohort (60% female, 57% white; S. Table1). The rate of CeD varied by ethnicity in the AoURP: 1.2% in white individuals, 0.4% in Hispanic individuals, and 0.2% in black individuals (S. Table 1). A clinical diagnosis of CeD typically requires serological and histological evidence(7,30). Serological data were unavailable for the majority of participants, 991 CeD participants have at least one available measurement (tTG-IgA, DGP-IgA, or DGP-IgG). Histological data concerning villous atrophy or crypt hyperplasia were unavailable.

### CeD patients with compatible HLA-DQ genotypes as primary analytic cohort

Among 3,481 individuals with documented or self-reported CeD, the HLA-DQ2.5 haplotype was enriched in participants with EHR-confirmed or self-reported CeD compared to non-CeD controls (both 43% vs. 23%, S. Fig. 2). No significant differences were observed in sex at birth, genetic ancestry, or HLA-DQ genotype distribution between the two CeD groups. Therefore, we included all 2,899 (83.3%) participants with one of the four well-established HLA-DQ risk haplotypes, and 14,495 propensity-matched non-CeD controls were included for further analysis (Fig. 1A-B, S.Fig.3). In this CeD cohort, 1904 had an ICD-coded CeD diagnosis in their EHRs, 1,666 self-reported CeD in surveys, and 771 had CeD documented in both sources (Fig. 1C). 333 individuals have positive serology. In seropositive and HLA-compatible cases as the primary analytic cohort, we found that 2,495 of the individuals with CeD were of European ancestry, 262 were of admixed American ancestry (89% self-identified as Hispanic), 108 were of African ancestry (71% self-identified as black), and 34 belonged to other ancestries (Fig. 1B).

**Figure 3.**
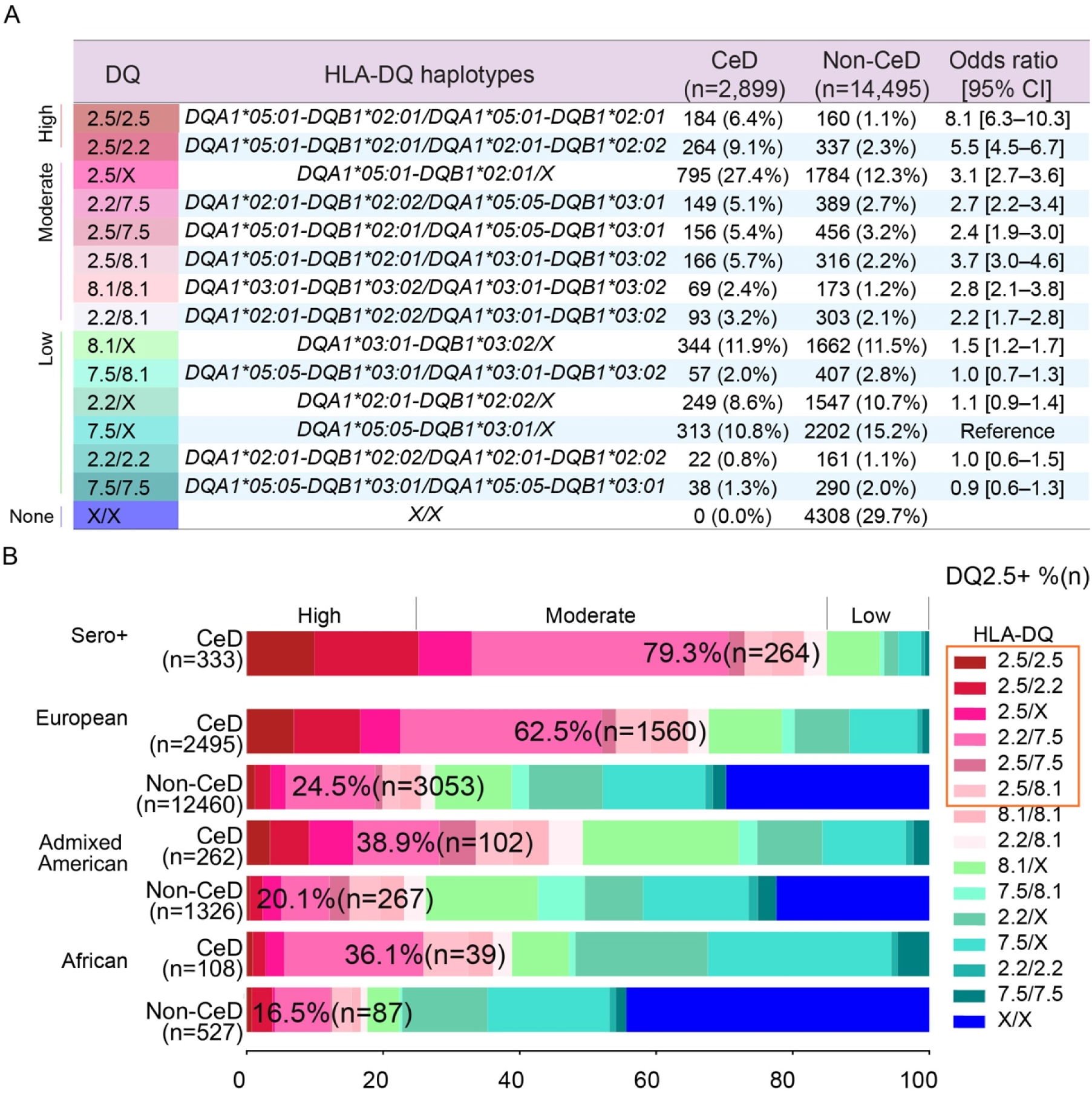
HLA-DQ genotype distribution and ancestry-specific effects in CeD. (A) Distribution of HLA-DQ genotypes based on the four known CeD-compatible HLA-DQ haplotypes among individuals with and without CeD. Genotypes were further categorized by risk level: high, moderate, low, or no risk (none). HLA-DQ 7.5/X was used as the reference, and p-values were FDR-adjusted. (B) HLA-DQ genotype distribution among seropositive CeD patients and across genetic ancestry groups (European, Admixed American, and African) for both the CeD and control cohorts. The frequency of individuals carrying HLA-DQ2.5, either in *cis* or *in trans*, is presented for each group.

### The prevalent AH8.1 ancestry haplotype in CeD

Only 13 of HLA alleles identified had a frequency that was more than 1% higher in CeD cases than in non-CeD controls (Fig. 2B, S.Table 4). Notably, an enrichment in *DQA1*05:01* and *DQB1*02:01*, which form the DQ2.5 heterodimer, was observed in CeD patients across all ancestries (Fig. 2B). The allele frequencies of *DQB1*02:01* varied by ancestry among individuals with CeD and non-CeD controls: 32.2% vs 11.6% in Europeans (OR=3.6, FDR p= 3.76E-257), 18.5% vs 8.1% in admixed Americans (OR=2.6, FDR p= 1.84E-10, Fig.2C). Among participants of African ancestry, the *DQB1*02:01* allele had a frequency of 15.7% (34/216) in CeD cases versus 8.1% (85/1,054) in non-CeD controls (OR = 2.1; FDR p = 0.005).

In non-CeD controls of European ancestry, the *DQA1*05:01-DQB1*02:01 (*DQ2.5) haplotype was in strong LD with A*01:01(A1), B*08:01(B8), C07:01(C7), and DRB1*03:01(DR3). B8 was linked to DQ2.5 in 74.2% of controls. This extended haplotype A1-B8-DR3-DQ2, known as ancestral haplotype 8.1 (AH8.1), has been implicated in multiple autoimmune diseases (14,31). This linkage was less pronounced in non-CeD controls of admixed American ancestry (42.1%, S.Table5). B8 was rare among individuals of African ancestry, with only 27 B8 alleles observed in the entire cohort (S.Table5). All observed B8 alleles were linked to *DQB1*02:01*, with an estimated B8–DQ2.5 linkage frequency of 9.7%–27.1% in non-CeD controls.

Next, we found that the frequency of the B8–DQ2.5 linkage was higher in individuals with CeD than in non-CeD controls across all ancestry groups: 78.2% vs 74.2% in Europeans (OR = 1.24, 95% CI 1.07–1.43, FDR p = 0.01), 54.6% vs 42.1% in admixed Americans (OR = 1.66, 95% CI 1.02–2.69, FDR p = 0.039; Table S2). For participants of African ancestry, B8–DQ2.5 linkage was more frequent in CeD cases (OR = 2.88, 95% CI 1.16–7.13, FDR p = 0.03, S.Table 6); however, the number of African CeD participants carrying DQ2.5 was small (n = 32, S.Table 7). Consequently, this estimate is based on fewer than 20 B8–DQ2.5–positive CeD cases, yields wide confidence intervals, and should be interpreted with caution.

Although DQ2.5 was observed across ancestries (11.6% in Europeans, 8.1% in admixed Americans, and 7.2% in Africans), the B8–DQ2.5 haplotype was much less frequent: 8.6% in Europeans, 3.4% in admixed Americans, and <1.9% in Africans (S. Table 5). Given the corresponding CeD prevalence estimates (1.2% in Europeans, 0.4% in admixed Americans, and 0.2% in Africans), these ancestry-specific LD patterns may contribute to differences in CeD prevalence across ancestries.

### Different clinical presentations associated with HLA-DQ risk genotypes

Our analysis of HLA-DQ genotypes showed that the highest risk of CeD was conferred by homozygosity for DQ2.5, followed by the DQ2.5/DQ2.2 genotype (Fig. 3A, S.Table 8). The combination of DQ2.5 and DQ2.2 was associated with a higher risk than DQ2.5/DQ7.5, consistent with a stronger additive effect of the DQB1*02 allele (32). The frequency of DQ2.5 carriers, either in *cis* (DQ2.5) or in *trans* (DQ2.2/7.5), was notably higher in CeD patients of European ancestry (62.5% vs 24.5%, Fig 3B), but this difference was less pronounced in those of American (38.9% vs 20.1%) or African ancestry (36.1% vs 16.7%, Fig. 3B). Genotypes such as homozygosity or heterozygosity for DQ2.2 or DQ7.5 were more frequent in controls than in CeD patients, consistent with their risks as permissive genotypes (17,18,32–35). These results again suggest while individuals of different ancestry having the DQ2.5 heterodimer, their risk for celiac disease is different, consistent with the observation that ancestry-specific B8-DQ2.5 LD patterns may contribute to differences in CeD prevalence. We then analyzed laboratory test results and comorbid conditions for the individuals for whom data were available. Based on the distribution of HLA-DQ genotypes in CeD cases and controls, we categorized participants into three risk groups: high-, moderate-, and low-risk (Fig. 3A). Serum autoantibody levels can decline significantly after the introduction of a GFD (36). We therefore used the highest recorded value for each antibody (tTG-IgA, DGP-IgA, and DGP-IgG) when multiple measurements were available for a participant. Individuals with high-or moderate-risk HLA-DQ genotypes had significantly higher mean levels of these CeD-specific antibodies than those with low-risk genotypes, highlighting a strong correlation between HLA-DQ genotype and serological markers of CeD (Fig. 4A). Interestingly, individuals with low-risk genotypes had fewer CeD-related visits documented in their EHRs (Fig. 4A). We compared clinical characteristics between CeD patients in three risk groups. High-risk CeD patients were more likely to have type 1 diabetes (T1D) and a family history of CeD, whereas those in the low-risk group more frequently presented with symptoms such as irritable bowel syndrome (IBS), diarrhea, or migraines, features more consistent with functional gastrointestinal disorders (Fig. 4B, S.Table 9). These findings suggest that, in some cases, the CeD diagnosis code may have been added to the EHR as part of diagnostic investigations rather than as a confirmed diagnosis. In summary, the high-and moderate-HLA-DQ risk genotypes are strongly associated with the presence of CeD-specific autoantibodies. Individuals with low-risk genotypes are more likely to display symptoms of functional gastrointestinal disorders, such as IBS, rather than true CeD.

**Figure 4.**
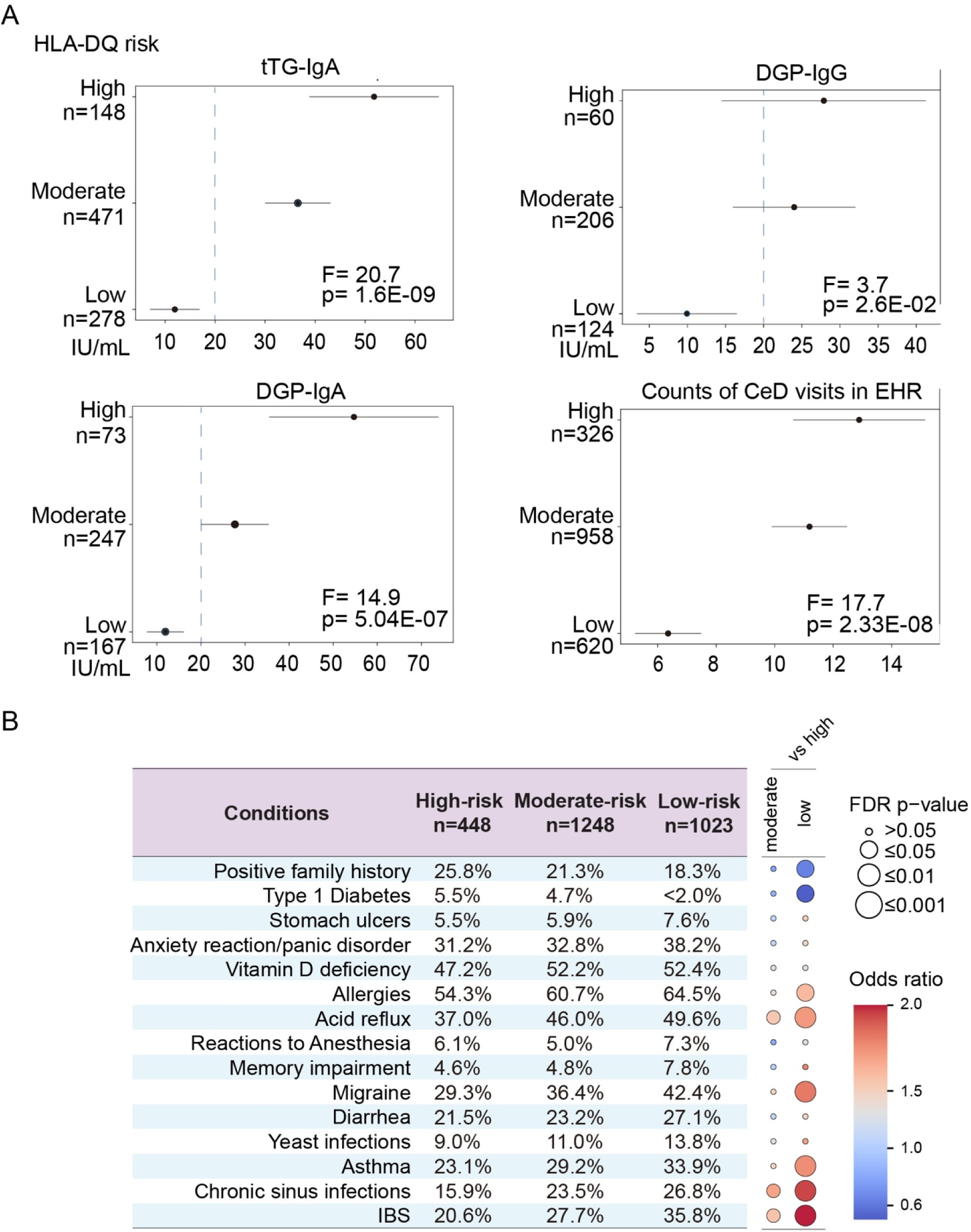
The frequencies of serological markers and comorbid clinical conditions differ between HLA-DQ genotypes in individuals with CeD (A) ANOVA was performed to compare differences in serological markers (tTG-IgA, DGP-IgA, DGP-IgG) and the number of clinical events classified as CeD recorded in the EHRs across HLA-DQ genotype categories. Tukey’s honestly significant difference post-hoc test was then performed to obtain mean values (black dots) and 95% confidence intervals (lines) for each group. Blue dotted lines indicate an arbitrary threshold value of 20 IU/mL. Visit counts represent unique encounters documented in the EHR on distinct dates. (D) Frequency of comorbid conditions across HLA-DQ risk categories in CeD patients, grouped into’high-risk’, ‘moderate-risk’ and’low risk’. A bubble plot displays the odds ratios and FDR adjusted p-values, using the high-risk group as the reference.

### Performance of a polygenic risk score (PRS) for predicting celiac disease across ancestries

A PRS derived from published GWAS and UK Biobank data, combining HLA-DQ genotypes with 38 non-HLA SNPs, effectively identifies individuals with CeD of European ancestry (17,18,32–35). We evaluated this log-additive PRS model in our cohort and confirmed that it could discriminate between 333 seropositive CeD cases and 676 non-CeD controls, achieving an area under the curve (AUC) of 0.86 (Fig. 5A-D). However, the model’s performance declined when applied to the full cohort or to individual ancestry groups. The median PRS in seropositive CeD cases was approximately 5.07, consistent with previous findings (32). PRS scores were significantly higher in CeD patients of European ancestry, intermediate in those of American ancestry, and discriminated poorly between cases and controls of African ancestry (Fig. 5B). Individuals of African ancestry had the lowest mean PRS scores, consistent with the observed lower risk of developing CeD in this population(B-D). Using the threshold that optimized prediction accuracy, we found that 33% of European CeD patients and 15% of seropositive CeD cases did not exceed the PRS cutoff. In addition, 26% of non-CeD European controls had PRS scores above the threshold, which may be partly explained by the presence of undiagnosed CeD among some controls. These findings suggest that, although the current PRS performs well in those with seropositive CeD (Fig 3D), the genetic risk score varies significantly in different populations, further work is needed for understanding the genetic risks for individuals of non-European ancestry.

**Figure 5.**
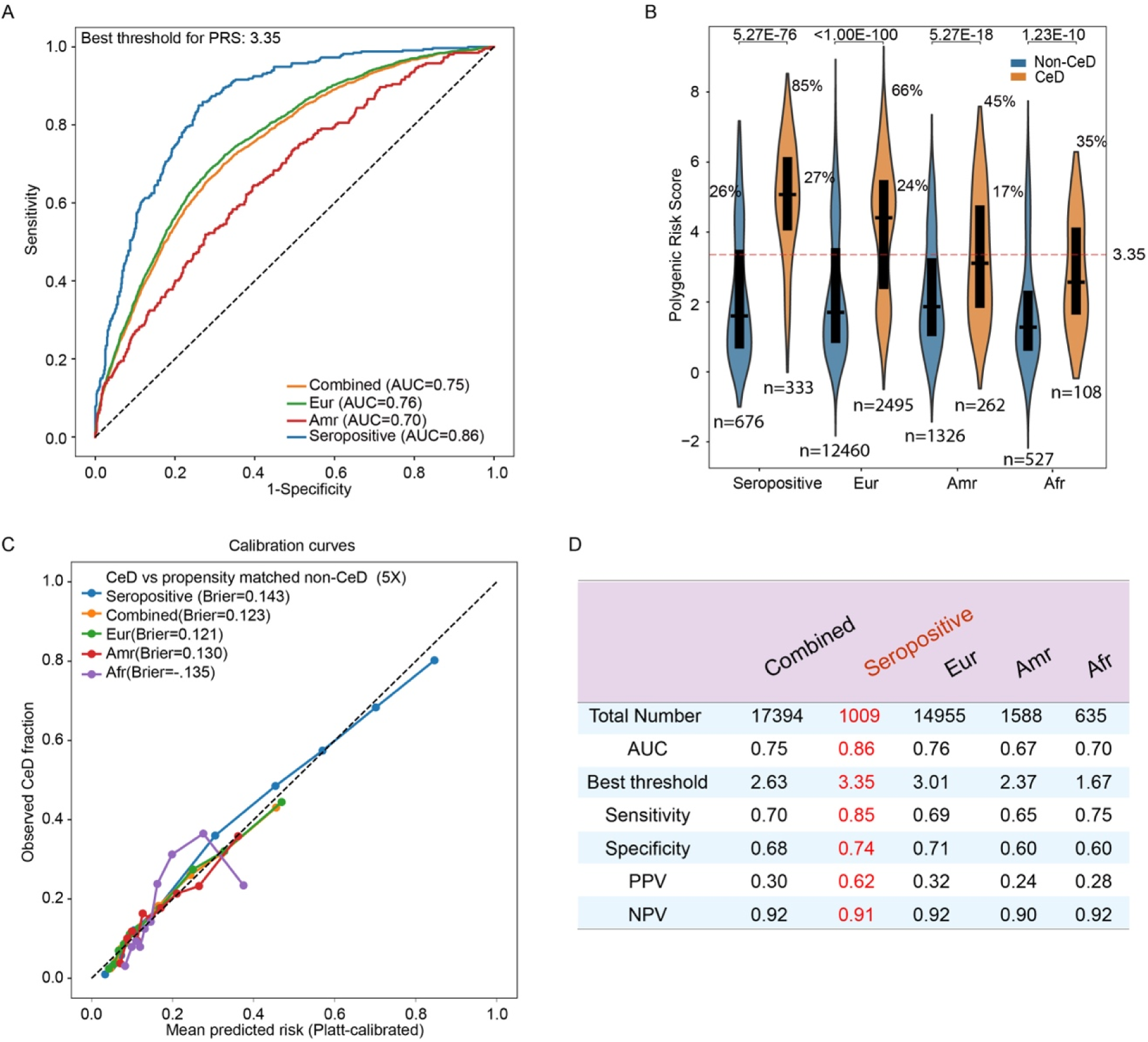
Evaluation of a reported PRS prediction model in participants from the *All of Us* project with CeD participants (A) ROC curves showing the performance of the PRS for distinguishing CeD cases from controls. The green line represents the ROC curve obtained with seropositive CeD cases considered positive and their matched non-CeD controls considered negative. PRS performance is evaluated across different ancestry subgroups: European, admixed American, and the combined cohort (regardless of ancestry). (B) Violin plots comparing PRS distributions between CeD patients and matched controls. PRS scores from seropositive CeD patients or those with European (Eur), admixed American (Amr), or African (Afr) ancestry were compared to their respective matched non-CeD controls. Mean PRS values are indicated by white dots. The dotted line represents the optimal PRS threshold with the highest accuracy for distinguishing seropositive cases from non-CeD controls. Percentages reflect the proportion of individuals in each group with PRS values above this threshold. The black boxes represent the interquartile range (25th to 75th percentile) of PRS values. (C) Platt post hoc probability calibration was applied by fitting a logistic regression that maps the model’s raw score (logit) to calibrated event probabilities. Calibration performance is summarized by the Brier score (mean squared error between predicted probabilities and observed outcomes), with lower values indicating better probabilistic accuracy. Bins centered at higher mean predicted risk indicate that the model assigns higher average risk in that subgroup (e.g., mean predicted risk >0.8 in the seropositive group). (D) Performance of the PRS for predicting CeD across subgroups defined by genetic ancestry and serologic status.

### CeD prediction model combining clinical and genetic risk factors

We then developed a predictive model incorporating HLA-DQ genotypes and a total of 50 features, including CeD-associated comorbid conditions. We evaluated machine-learning approaches, including decision trees and logistic regression, using seropositive CeD patients as positive cases (S. Methods). Logistic regression identified seven independent predictors: high-or moderate-risk HLA-DQ genotype, family history of CeD, diarrhea, vitamin D deficiency, anemia, and hypothyroidism. This model achieved an AUC of 0.87 for CeD prediction (Fig. 6A). As a means of improving prediction further, we developed a composite log-additive AoU-CeD score that integrates family history, four clinical diagnoses, and PRS. This combined model outperformed the PRS alone. Using a threshold score of 4.17 to achieve 90% sensitivity for the detection of seropositive CeD, we assessed the performance of our model in patients lacking strong serological biomarkers. The threshold was reached for 77% of CeD patients of European ancestry, with lower proportions attained in the American and African ancestry groups (Fig. 6C).

**Figure 6.**
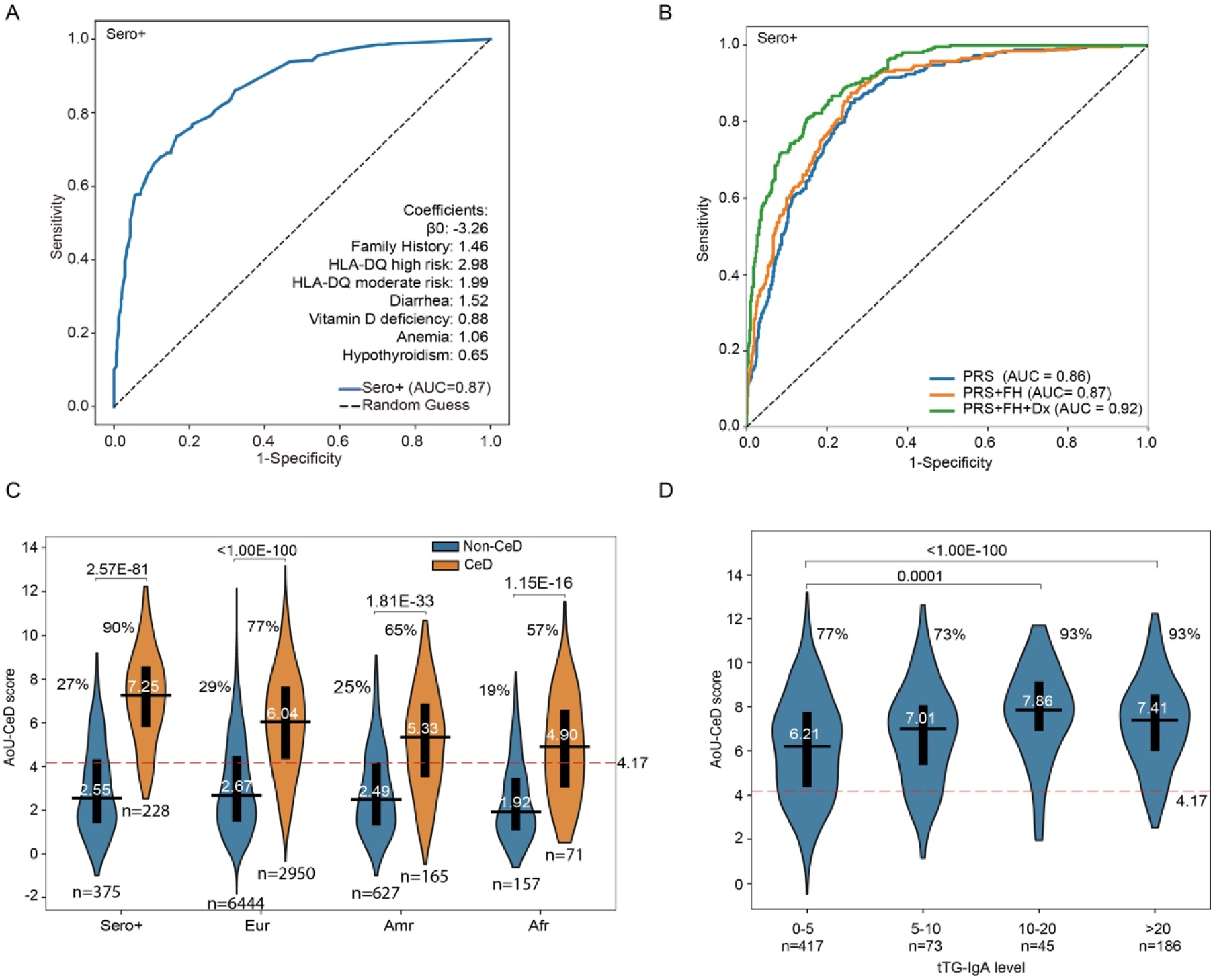
The AoU-CeD score combines clinical risk factors and PRS to improve CeD prediction over and above that achieved with the PRS alone (A) Logistic regression and machine-learning approaches were used to identify the variables that best predicted CeD. The input variables included sex, age, genetic ancestry, HLA-DQ genotype, and 42 clinical features, including symptoms, comorbid conditions and complications related to malabsorption. Seropositive CeD participants were used as positive cases. A ROC curve was plotted and the model coefficients for the top predictors are reported. (B) We evaluated the extent to which the incorporation of clinical data improved prediction over that achieved with the PRS alone, by testing models including family history (FH) and seven clinical diagnoses (Dx). ROC curves comparing three scoring methods: PRS alone, PRS + FH, and PRS + FH + Dx. Scores were calculated with a log-additive model, combining log-transformed values weighted by logistic regression coefficients. **(C)** Violin plots showing AoU-CeD score distributions in seropositive CeD cases versus matched non-CeD controls, using a threshold capturing 90% of seropositive CeD patients. The AoU-CeD score was then applied to CeD patients without high levels of serological markers (>20 IU/mL), stratified by ancestry. **(D)** Validation of the AoU-CeD score in CeD patients from *All of Us* with available but variable tTG-IgA values recorded in their EHRs.

We then validated the model by investigating its relationship to tTG-IgA levels. We found that 186 CeD patients had tTG-IgA levels >20 IU/mL, and 45 had levels between 10–20 IU/mL. Among those with tTG-IgA levels >10 IU/mL, 93% surpassed the predictive threshold, versus 77% with levels <5 IU/mL. These results support the utility of this model for stratifying seropositive CeD cases.

## Discussion

A major advantage of the *All of Us* dataset is its enormous volume of EHRs, genomic and survey data, including the most comprehensive set of information available for populations historically underrepresented in medical research. This enabled us to study CeD patients from diverse ancestries in this first study to investigate extensive genetic data from patients from African and admixed American ancestry.

The *All of Us* data confirm that CeD diagnosis is positively associated with sex (more frequent in women) and socioeconomic status but negatively associated with black or Hispanic ethnicity (2,28,37). Clinical epidemiological studies, based principally on the serological marker tTG-IgA, have shown that the prevalence of seropositive CeD varies across countries and racial/ethnic group. The prevalence of seropositivity ranges from 0.3% in Germany, to 2.4% in Finland (38), and 2% in Norway (HUNT study) (38), with an overall estimated prevalence of 1% in Europe, 0.7% in the United States, 0.5% in South America, 0.34% to 1.1% in Africa, and 0.36% in China (2). In the All of Us cohort, CeD prevalence varied by self-reported race/ethnicity—1.2% in White individuals, 0.4% in Hispanic individuals, and 0.2% in Black individuals—which is consistent with findings from the National Health and Nutrition Examination Survey (NHANES) 2009–2012 (39).

In NHANES, based on tTG and endomysial (EMA) IgA antibody testing in 14,701 participants, CeD prevalence was highest among White individuals (1.08%) and substantially lower among Mexican Americans (0.23%), other Hispanics (0.38%), and Black individuals (0.22%) (39).

This raises the question as to what drives ancestry-specific variation in seropositivity rates. HLA-DQ2.5 frequency is only modestly lower across ancestries—11.6% in Europeans, 8.1% in individuals of admixed American ancestry, and 7.2% in Africans—yet CeD prevalence in Africans and admixed Americans appears disproportionately lower than would be expected based on DQ2.5 carriage alone. One possible explanation lies in the LD patterns of HLA alleles. The high-risk haplotype (B8: C7: DR3: DQ2.5) is present in 72.3% of DQ2.5-positive individuals of European ancestry, but in less than 20% of those of African ancestry. Our findings suggest a potential explanation: individuals of African ancestry may carry the canonical DQ2.5 risk heterodimer, yet reduced linkage to B8—or to B8-associated genetic factors—could influence the likelihood or magnitude of a tTG-IgA production. For example, the *MICA*5.*1 allele is in strong linkage disequilibrium with B8 and may modify DQ2.5-mediated antigen presentation (40). An additional possibility is that patients with CeD of non-European ancestry may not mount tTG-IgA responses comparable to those of European ancestry. In NHANES, a discordant pattern between tTG-IgA and EMA-IgA was observed among participants of African ancestry(39), and in an Alabama-based study (in which 26% of residents were of African ancestry), tTG-IgA was more frequently reported as negative (41). Undermeasurement and seronegative disease have therefore been proposed as alternative explanations for the apparently lower frequency of CeD among individuals of African descent (42). Finally, tTG-IgA quantification is not fully standardized: upper limits of normal vary substantially across laboratories, commercial assays, and serum dilution protocols, complicating the definition of reliable cutoff values(36,43,44). This inter-assay variability may contribute to false-negative serologic results and may disproportionately hinder accurate CeD ascertainment in individuals of non-European ancestry.

HLA-DQ testing is included in both the American College of Gastroenterology (ACG) and American Gastroenterological Association (AGA) guidelines for the diagnostic evaluation of CeD (15,16). According to these guidelines, genetic testing for CeD-compatible HLA haplotypes is not necessary in all cases but may be useful in specific situations, such as cases in which there is a discrepancy between serology and histology results, or in individuals who have already switched to a GFD before evaluation. A negative result of HLA-DQ genotypes effectively rules out CeD. The presence of a CeD-compatible haplotype may support gluten challenge in appropriate clinical contexts. We found that 17% of individuals with a CeD diagnosis did not carry any of the four historically defined CeD-compatible HLA-DQ haplotypes. Conversely, about 70% of non-CeD controls carried these haplotypes. Our data suggest that CeD is frequently misdiagnosed in the United States; in particular, many patients lack serologic evidence of CeD and may carry low-risk HLA-DQ genotypes.

Can predictive models improve CeD risk stratification? Prediction models for CeD have been developed based on symptoms, comorbid conditions (45,46), or polygenic risk scores (32). Our preliminary prediction model supports a case-finding approach, with effective risk stratification and targeted identification of high-risk individuals. A cost-effectiveness analysis showed that testing children with a pre-test probability ≥10% by both HLA typing and tTG-IgA was the most effective approach (47). HLA testing before tTG-IgA testing was the most cost-effective approach for predicting CeD (48). As genome sequencing becomes more affordable, it will become possible to use HLA risk stratification to optimize population-level CeD screening further (44). Further studies are required to assess whether AoU-CeD score combining clinical factors and PRS can aid in clinical management in independent cohorts. For example, can the score identify patients with an ambiguous diagnosis who are most likely to benefit from a gluten challenge, and how does its predictive performance compare with established endpoints such as seropositivity and histologic response? In addition, is the score concordant with tTG-IgA levels when screening first-degree relatives?

However, there are important limitations in our study. Key clinical details are missing for many participants, such as pathology reports for endoscopy examinations and dietary information, including whether patients are already following a gluten-free diet or how long they have been on it. Future studies incorporating dietary factors would be highly valuable. Other limitations include incomplete survey participation and limited availability of EHRs for review; moreover, only a small subset of patients had serologic measurements of tTG and DGP antibodies. Another concern is the potential inaccuracy of EHR-derived phenotypes: diagnostic codes may be assigned primarily for billing purposes rather than to reflect a confirmed diagnosis. Some patients labeled with CeD may have been diagnosed with seronegative or potential CeD, rather than seropositive and histologically confirmed disease. We observed that chronic conditions associated with functional gastrointestinal disorders were prevalent, affecting 35.8% of individuals with low-risk *HLA-DQ* genotypes. This suggests that a subset of patients labeled with CeD may actually have other conditions, such as non-celiac gluten sensitivity(49) or functional gastrointestinal disorders(48). These findings highlight the need for more stringent criteria for identifying CeD patients in future genetic studies, particularly among patients with non-European ancestry.

In conclusion, genetic risks can differ significantly across individuals of diverse ancestry, and integrating polygenic risk scores with family history, symptoms, and comorbid conditions provides a promising approach for improving the screening and diagnosis of celiac disease.

## Data Availability

The data and code used in this study are available as a shared workspace to registered researchers of the All of Us Researcher Workbench. For information about access, please visit https://www.researchallofus.org/.

## Notes

**Funding** This work was supported by the National Institute of Diabetes and Digestive and Kidney Diseases of the National Institutes of Health (K08DK128631 and R03DK144282 to X.-F.K.) and by the Disease-Oriented Clinical Scholars Program at the University of Texas Southwestern Medical Center.

**Acknowledgments** The *All of Us* research program is supported by the National Institutes of Health, Office of the Director: Regional Medical Centers: 1 OT2 OD026549; 1 OT2 OD026554; 1 OT2 OD026557; 1OT2 OD026556; 1 OT2 OD026550; 1 OT2 OD 026552; 1 OT2 OD026553; 1 OT2 OD026548; 1OT2 OD026551; 1 OT2 OD026555; IAA #: AOD 16037; Federally Qualified Health Centers: HHSN 263201600085U; Data and Research Center: 5 U2C OD023196; Biobank: 1 U24OD023121; The Participant Center: U24 OD023176; Participant Technology Systems Center: 1U24 OD023163; Communications and Engagement: 3 OT2 OD023205; 3 OT2 OD023206; and Community Partners: 1 OT2 OD025277; 3 OT2 OD025315; 1 OT2 OD025337; 1 OT2OD025276. The *All of Us* Research Program would not be possible without the partnership established with its participants.

### Competing Interest Statement

The authors have declared no competing interest.

### Funding Statement

The All of Us research program is supported by the National Institutes of Health, Office of the Director: Regional Medical Centers: 1 OT2 OD026549; 1 OT2 OD026554; 1 OT2 OD026557; 1OT2 OD026556; 1 OT2 OD026550; 1 OT2 OD 026552; 1 OT2 OD026553; 1 OT2 OD026548; 1OT2 OD026551; 1 OT2 OD026555; IAA #: AOD 16037; Federally Qualified Health Centers: HHSN 263201600085U; Data and Research Center: 5 U2C OD023196; Biobank: 1 U24OD023121; The Participant Center: U24 OD023176; Participant Technology Systems Center: 1U24 OD023163; Communications and Engagement: 3 OT2 OD023205; 3 OT2 OD023206; and Community Partners: 1 OT2 OD025277; 3 OT2 OD025315; 1 OT2 OD025337; 1 OT2OD025276. The All of Us Research Program would not be possible without the partnership of its participants. This work was supported by the National Institute of Diabetes and Digestive and Kidney Diseases of the National Institutes of Health (K08DK128631 and R03DK144282 to X.-F.K.) and by the Disease-Oriented Clinical Scholars Program at the University of Texas Southwestern Medical Center.

### Author Declarations

All data collection and analysis involving human participants were approved by the Ethics Committee/Institutional Review Board (IRB) of the All of Us Research Program (AoU IRB Protocol Number: 2021-02-TN-001).

### Summary of Updates

In this revision, we used the updated, curated data repository from All of Us C2024Q3R4, which includes a larger number of participants with available genetic data. Second, we excluded participants with celiac disease who do not carry HLA-DQ risk genotypes from the analysis. Third, we incorporated a polygenic risk score to discriminate between cases and controls.

