## Supplementary methods and figures for "Linkage between HLA-B8 and HLA-DQ2.5 Contributes to Ancestry-Dependent Genetic Risk for Celiac Disease"

**Supplementary Figures:**

**Supplementary Figure 1: HLA and concordance analysis across three HLA typing methods.** We then tested three strategies, including HIBAG, HLA*LA, and tagSNPs. We found the inconsistency arose from *HLA-DQA1* imputation. Both HIBAG and HLA*LA make errors in calling HLA-DQA1, particularly DQA1*0501 and DQA1*0505, however there was a high level of consistency for *HLA-DQB1* and *DRB1*. Combining tagSNPs and HIBAG methods, we were able to impute HLA-DQ genotype with a high degree of consistency, relying principally on the ancestry specific LD between *HLA-DQB1* and *DRB1* for the assignment of haplotypes.

(A) Venn diagrams of HLA typing results for HLA-LA and HIBAG. Beige: total matched, salmon: HIBAG, green: HLA-LA. (B) Venn diagrams of HLA-DQ genotypes for HIBAG, HLA-LA, and tag-SNP. Salmon: tag-SNP, green: HIBAG, dark blue: HLA-LA: bright blue: tag-SNP and HLA-LA overlap, beige: tag-SNP and HIBAG overlap, sky-blue: HLA-LA and HIBAG overlap, grayish purple: total matched. HIBAG: HLA genotype imputation with attribute bagging, HLA-LA: HLA-linear alignment.

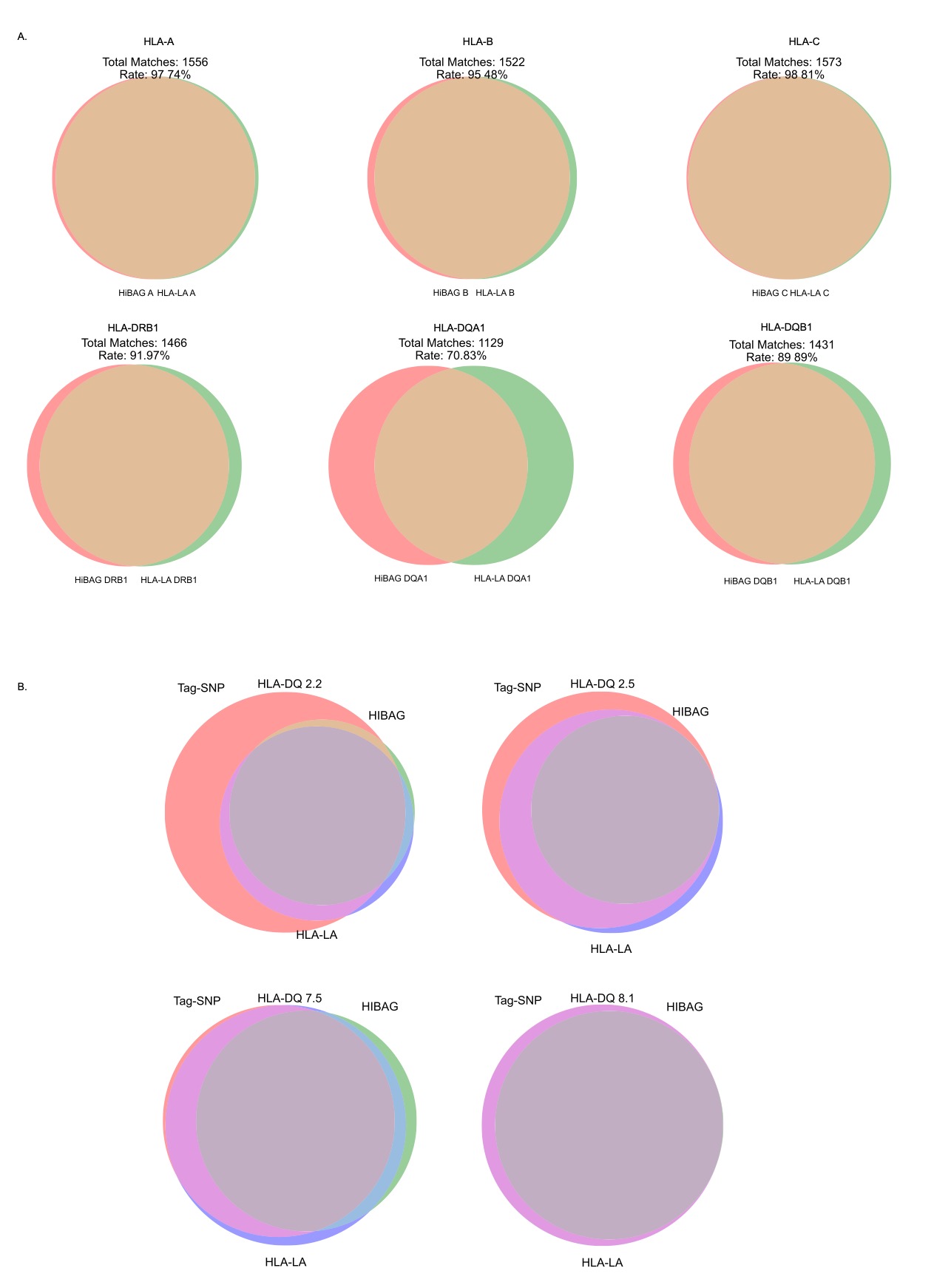

**Supplementary Figure 2:** Comparison of multiple variables between CeD participants identified by EHR and Self-Reported.

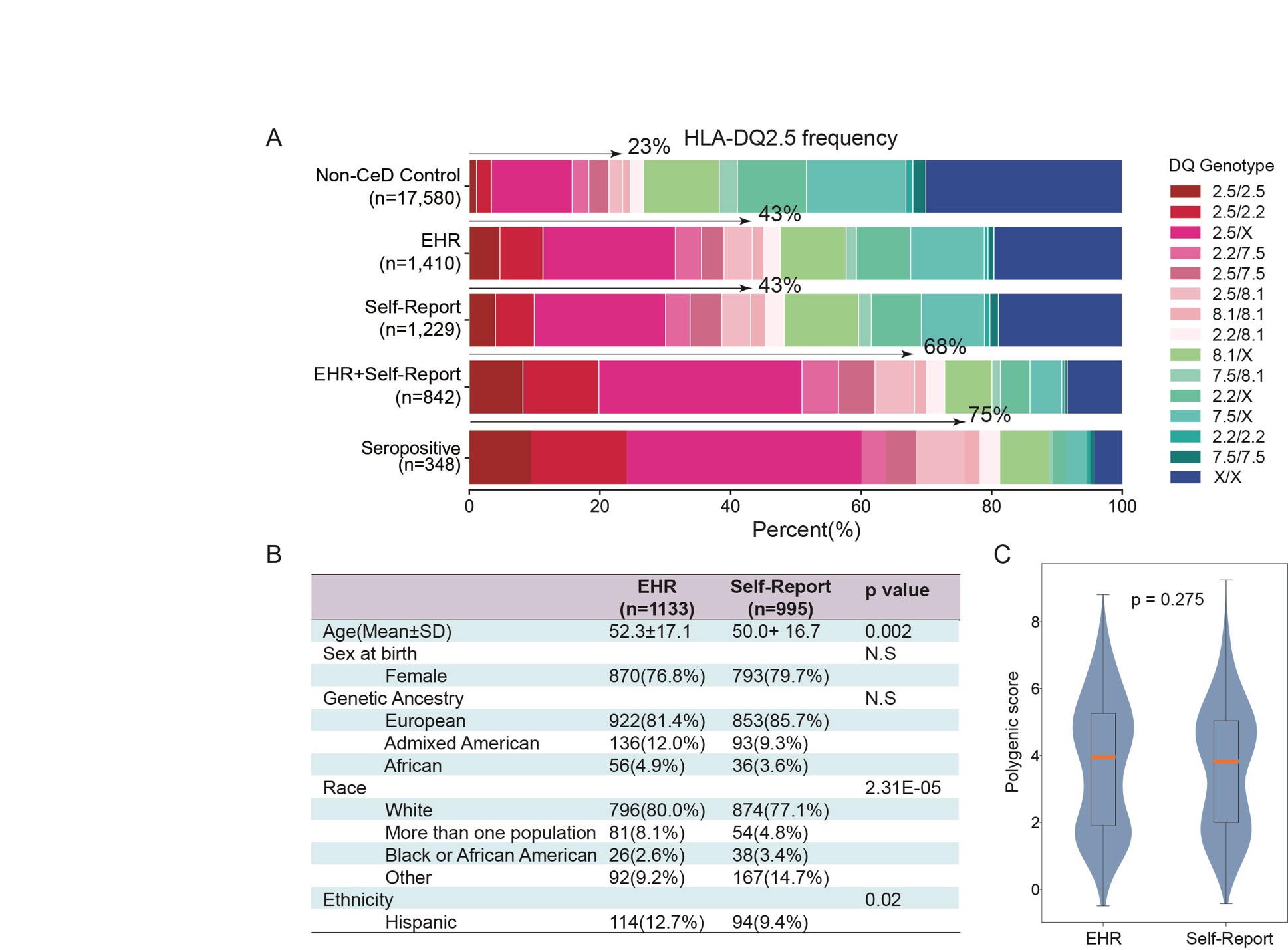

(A) Stacked plot showing the distribution of HLA-DQ genotypes among CeD participants from the five groups before removing those without compatible HLA-DQ haplotypes.

(B) Comparison of sex, genetic ancestry, race, and ethnicity among CeD participants with compatible HLA-DQ genotypes. SD: standard deviation.

(C) Comparison of polygenic risk scores between participants identified by EHRs only and those identified by self-report only.

**Supplementary Methods**

**Workflow for developing the logistic regression model for CeD classification**

We built a supervised classification model to distinguish CeD from non-CeD controls, using seropositive CeD cases for model training. Predictors included demographic variables, clinical histories, and the ordered HLA-DQ impact label (Low/Moderate/High). Specifically, we included sex at birth, age, two numeric symptom scores (“Overall Health: Average Pain 7 Days” and “PROMIS-MH”), a set of binary clinical variables, and the HLA-DQ impact label. The impact label was encoded with Low as the reference category, generating two dummy variables (Moderate and High).

Numeric predictors were coerced to numeric values and missing values were imputed using the median of the observed distribution. Binary predictors were harmonized by mapping yes/no-like values to 1/0 and setting missing/unknown values to 0. After encoding, predictors with all missing values or zero variance were removed. To reduce instability in model fitting and mitigate quasi- or perfect separation, we excluded extremely sparse binary predictors where either the number of positives or negatives was <5. We also removed duplicate predictors and pruned near-perfectly correlated predictors by dropping one variable from any pair with an absolute Pearson correlation >0.99.

Feature selection was performed using L1-penalized logistic regression (lasso) implemented in scikit-learn. Predictors were standardized within a pipeline, and we used class_weight = “balanced” to account for class imbalance. We tuned the regularization strength across a log-spaced grid of inverse-penalty parameters (C) using stratified K-fold cross-validation (5 folds) and ROC AUC as the performance metric. The final model was selected using a parsimony criterion (one–standard-error rule), choosing the sparsest model whose mean AUC was within one standard deviation of the best-performing model. Predictors with non-zero L1 coefficients in the chosen model were retained as the final feature set.

After feature selection, we refit an unpenalized logistic regression using statsmodels (Logit) with an intercept term, using the L-BFGS optimizer for numerical stability. For each retained predictor, we estimated the fitted regression coefficients (log-odds) from the refit logistic model and converted them to odds ratios (ORs) by exponentiation (OR = exp[β]). Predicted probabilities were computed using a numerically stable logistic function, and model discrimination was summarized by the area under the ROC curve (AUC).

**List of all 50 features used to train the logistic regression model and derive the clinical risk coefficients.**

I. Patient-Reported Experience & Quality of Life (2 variables)

1. PROMIS total score

2. Overall health – average pain in past 7 days

II. Clinical History (42 variables)

3. Diarrhea

4. Crohn's disease

5. Acid reflux

6. Allergies

7. An eating disorder

8. Anxiety reaction or panic disorder

9. Asthma

10. Autism spectrum disorder

11. Chronic fatigue

12. Depression

13. Endometriosis

14. Fibroids

15. Fibromyalgia

16. Hyperthyroidism

17. Memory loss or impairment

18. Neuropathy

19. Osteoporosis

20. Peptic ulcers

21. Polycystic ovarian syndrome

22. Restless leg syndrome

23. Skin conditions (e.g., eczema, psoriasis)

24. Spine, muscle, or bone disorders (non-cancer)

25. Type 1 diabetes

26. Ulcerative colitis

27. Vitamin B deficiency

28. Chronic sinus infections

29. Lyme disease

30. Recurrent urinary tract infections

31. Recurrent yeast infections

32. Vitamin deficiency

33. Irritable bowel syndrome (IBS)

34. Hypothyroidism

35. Vitamin D deficiency

36. Anemia

37. Migraine

38. Thyroiditis

39. Reactions to anesthesia (e.g., hyperthermia)

40. Systemic lupus

41. Functional digestive disorders

42. Other disorders of the stomach and duodenum

43. Noninfectious gastroenteritis

44. Symptoms involving the digestive system

III. Demographic & Genetic Features (5 variables)

45. Sex

46. Age

47. Predicted genetic ancestry

48. HLA-DQ high-risk genotype

49. HLA-DQ moderate-risk genotype

IV. Behavioral & Family History (1 variable)

50. Family history of celiac disease
